# Preventable suicides involving medicines before the covid-19 pandemic: a systematic case series of coroners’ reports in England and Wales

**DOI:** 10.1101/2023.07.02.23292130

**Authors:** Grace Anthony, Jeffrey K. Aronson, Richard Brittain, Carl Heneghan, Georgia C. Richards

## Abstract

**Background:** Over 5000 suicides are registered in England and Wales each year. Coroners’ Prevention of Future Deaths reports (PFDs) share concerns to promote actions to reduce the risks of similar deaths.

**Aims:** To systematically review coroners’ PFDs involving suicides in which a medicine caused or contributed to the death, to identify lessons for suicide prevention.

**Methods:** 3037 PFDs were screened for eligibility between July 2013 and December 2019. Following data extraction, descriptive statistics and content analysis were performed to assess coroners’ concerns, the recipients, and the response rates to reports.

**Results:** There were 734 suicide-related PFDs, with 100 reporting a medicine. Opioids (40%) were the most common class involved in suicide-related PFDs, followed by antidepressants (30%). There was wide geographical variation in the writing of PFDs; coroners in Manchester wrote the most (18%). Coroners expressed 237 concerns; the most common were procedural inadequacies (14%, n=32), inadequate documentation and communication (10%, n=22), and inappropriate prescription access (9%, n=21). 203 recipients received these PFDs, most being sent to NHS trusts (31%), clinical commissioning groups (10%), and general practices (10%), of which only 58% responded to the coroner.

**Conclusions:** Concerns raised by coroners in suicide-related PFDs involving medicines highlight essential gaps in care that require actions from the Government, health services, and prescribers. To aid suicide prevention, PFDs should be disseminated nationally, and responses should be enforced so that actions are taken to prevent suicides.

**Study protocol registration:** https://doi.org/10.17605/OSF.IO/EXJK3

## Introduction

Suicide is a leading cause of preventable deaths, involving over 700,000 deaths every year globally (1). In England and Wales, 5691 suicides occurred in 2019, a 12% increase since 2000 (2). In 2021, there were 5583 registered suicides in England and Wales (3). Various factors are associated with the risk of suicide, including a previous attempt, self-harm, bereavement, mental ill-health, social isolation, and recent adverse life events (1, 4, 5).

Legislation to reduce pack sizes of over-the-counter (OTC) paracetamol has been effective in reducing deaths, with a 35% reduction in paracetamol poisonings between 1998 and 2009 (6). The opioid, co-proxamol, was removed from UK markets in 2007 owing to its toxicity in suicide, with an estimated 500 lives saved between 2010 and 2015 (7). Other opioids, such as codeine, are still available OTC in the UK with large volumes being sold (8).

Coroners in England and Wales have a duty to communicate a death when they believe that actions should be taken to prevent future deaths (9). These reports, called Prevention of Future Deaths reports (PFD) (10–12), are sent to individuals or organisations with the power to act. Once received, recipients must respond to the coroner within 56 days, outlining actions taken or proposed. However, there is no mechanism to enforce responses or to analyse, monitor, or learn from PFDs (13).

Previous studies have assessed suicide-related PFDs (13–15), but these have focused on specific issues, including claims against the NHS. The Guardian reviewed 500 PFDs in 2018 to identify deaths related to mental illness (16, 17). A systematic case series of PFDs involving medicines was also conducted in 2013-2022, identifying opioids and antidepressants as the leading medicines involved in all preventable deaths (18). The ONS has published an analysis of 164 suicide-related PFDs, covering 2021 and 2022 alone (19). However, no research to date has investigated PFDs involving medicines and suicides before the SARS-CoV-2 pandemic. We therefore aimed to assess all available PFDs detailing deaths by suicide in which a medicine caused or contributed to the death. Our primary objectives were to discover: 1) the medicines involved; 2) the concerns raised by coroners; 3) the recipients to whom PFDs were addressed; and 4) the response rates of addressees.

## Methods

We designed a retrospective case series and pre-registered the study protocol on an open repository (https://doi.org/10.17605/OSF.IO/EXJK3).

### Data sources

PFDs have been openly available on the Courts and Tribunals Judiciary website since 2013 (20). We used web scraping to automatically collect all published PFDs to create a database and build the Preventable Deaths Tracker (https://preventabledeathstracker.net/) (21). The code to create the web scraper is available on GitHub and has been previously described (22, 23). The database contains the case reference number; the date of the report; the name of the deceased; the coroner’s name; the coroner’s area; the category of death (defined by the Chief Coroner’s Office); to whom the report was sent; and the url of the Judiciary website. Population data for suicides in England and Wales corresponding to the dates of PFDs included in the analysis (2013–2019) were obtained from the Office for National Statistics (ONS) (2).

### Eligibility of PFDs

There were 3037 PFDs available at the time of the web scrape. PFDs were screened using a pre-defined algorithm (Supplement Figure S1), in which suicide was defined as intentional self-harm that led to or contributed to death or death of undetermined intent by injury or poisoning. This definition was widened compared with the ONS definition of suicide, because the standard of proof for reaching a conclusion of suicide was changed from a criminal to a civil one in 2018 (24). The ONS relies on the conclusion supplied by the coroner, which is influenced by factors such as the age and sex of the deceased, the suicide method, marital status, and the individual likelihood that the coroner will supply a narrative verdict (25–28). If there were discrepancies in eligibility, cases were discussed with the authorship team and a conclusion was reached.

Following the inclusion of all suicide-related PFDs, cases were reviewed for medicines. PFDs were included if a medicine was listed as a medical cause of death or was stated as contributing to the death by the coroner or was the cause of toxicity resulting in death. A medicine was defined as any prescription-only or over-the-counter medication. Cases were excluded if the drug or drugs were obtained illicitly, or if the suicide involved non-medical products (e.g. alcohol), or if the medicine was mentioned in the report, but there was no indication that it directly or indirectly contributed to the death.

### Data extraction

Following the screening for eligibility, one study author (GA) manually extracted data from the included PFDs into a predesigned Google Sheet. The data extracted, included:

1. the date of death,
2. the inquest start date and end date,
3. the Judiciary website categorisation,
4. the age of the deceased (if reported),
5. the sex of the deceased,
6. the location of the death (if noted),
7. the medicine that caused or contributed to the death,
8. how the medicines were obtained,
9. health service contacts before the death,
10. relevant medical, mental health, and social history,
11. concern(s) raised by the coroner,
12. those who responded to the PFD, and
13. the date on which the response was received.

### Data analysis

We descriptively analysed the extracted data using medians and interquartile ranges (IQR) for continuous variables and frequencies (percentages) for categorical variables. The types of medicines were categorised into drug classes using the British National Formulary (BNF). To examine geographical variation, we further grouped coroner areas into standard regions to create a choropleth map. We used content analysis with inductive logic to group concerns raised by coroners from included PFDs. Common themes and phrases were highlighted and drawn out, and frequencies were calculated using the total numbers of concerns as the denominators. We calculated a response rate to PFDs for each organisation as the proportion of reports to which a response was submitted over the total number received. In October 2020, responses were further classified as either early (delivered before 56 days), on-time (delivered on the 56th day), late (submitted after 56 days), or overdue (when no response was present on the Judiciary website).

### Software and data sharing

We used Google Sheets (29) to analyse the data, and Tableau to create graphs. The data and study materials are openly available via the Open Science Framework (OSF), and the web scraper code is available on Github (30). Datawrapper was used to create a choropleth map of published PFDs by regions of England and Wales (https://www.datawrapper.de).

### Protocol deviations

The registered study protocol was designed to analyse all suicide-related deaths reported in PFDs. However, the team decided to focus the main analysis on suicides involving medicines due to the increasing concerns of self-poisonings with medicines. We had also planned to analyse the text of the responses and submit freedom of information (FOI) requests to increase the number of responses to PFDs, but this analysis was not conducted due to resource constrains.

## Results

734 PFDs (n=735 deaths) involved a suicide between July 2013 and December 2019 (Supplement Figure S2). Of these suicide-related PFDs, 14% (n=100) reported that a medicine caused or contributed to the death. Over the 6.5-year study period, there was a median of 118 (IQR: 102-119) suicide-related PFDs each year and a median of 17 (IQR: 10-19) medicine-related suicides (Figure 1; Supplement Table S1).

**Figure 1:**
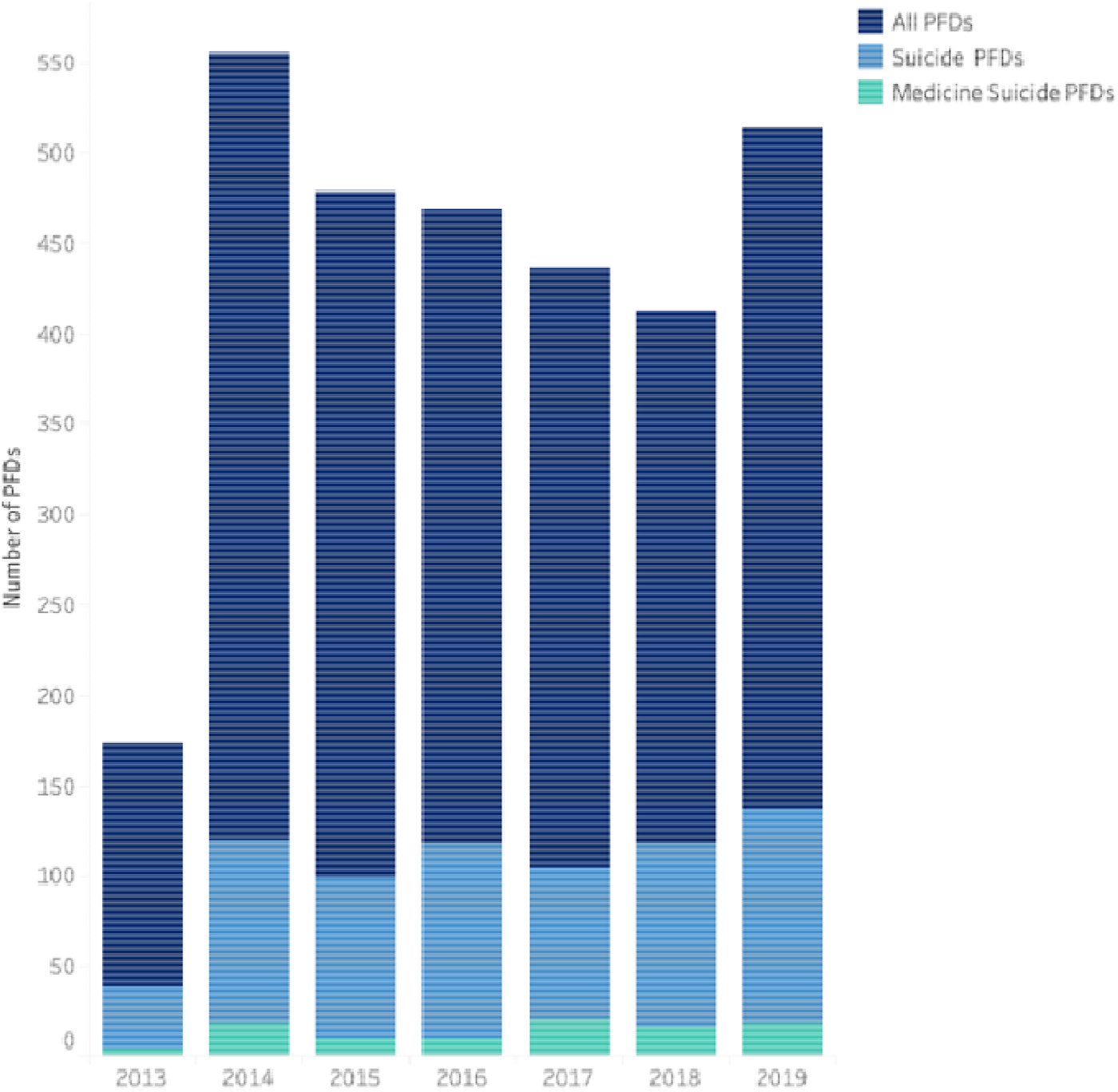
Trends of coroners’ Prevention of Future Deaths reports (PFDs) published between July 2013 and December 2019 involving suicides (light blue) and medicine-related suicides (green) compared with the total number of published PFDs (dark blue).

On the judiciary website PFDs are assigned one or more categories; of our 100 medicine suicide PFDs there were 48 Hospital (Clinical Procedures and medical management) related deaths, 20 Community Healthcare and Emergency Services related deaths, 19 Suicide (2015) related deaths and 17 Alcohol, drug and medication related deaths (Supplement Figure S3). In the 100 PFDs involving medicines and suicides, 46 unique medicines were identified, involving a total of 152 medicines. Morphine (15%) was the most often reported, followed by paracetamol (12%) and zopiclone (10%) (Figure 2). Using BNF drug classes, opioids (n=40) caused or contributed to the most medicine-related suicides, followed by anti-depressants (n=30) (Supplement Table S2). In 16 PFDs the type of medicine was unspecified.

**Figure 2.**
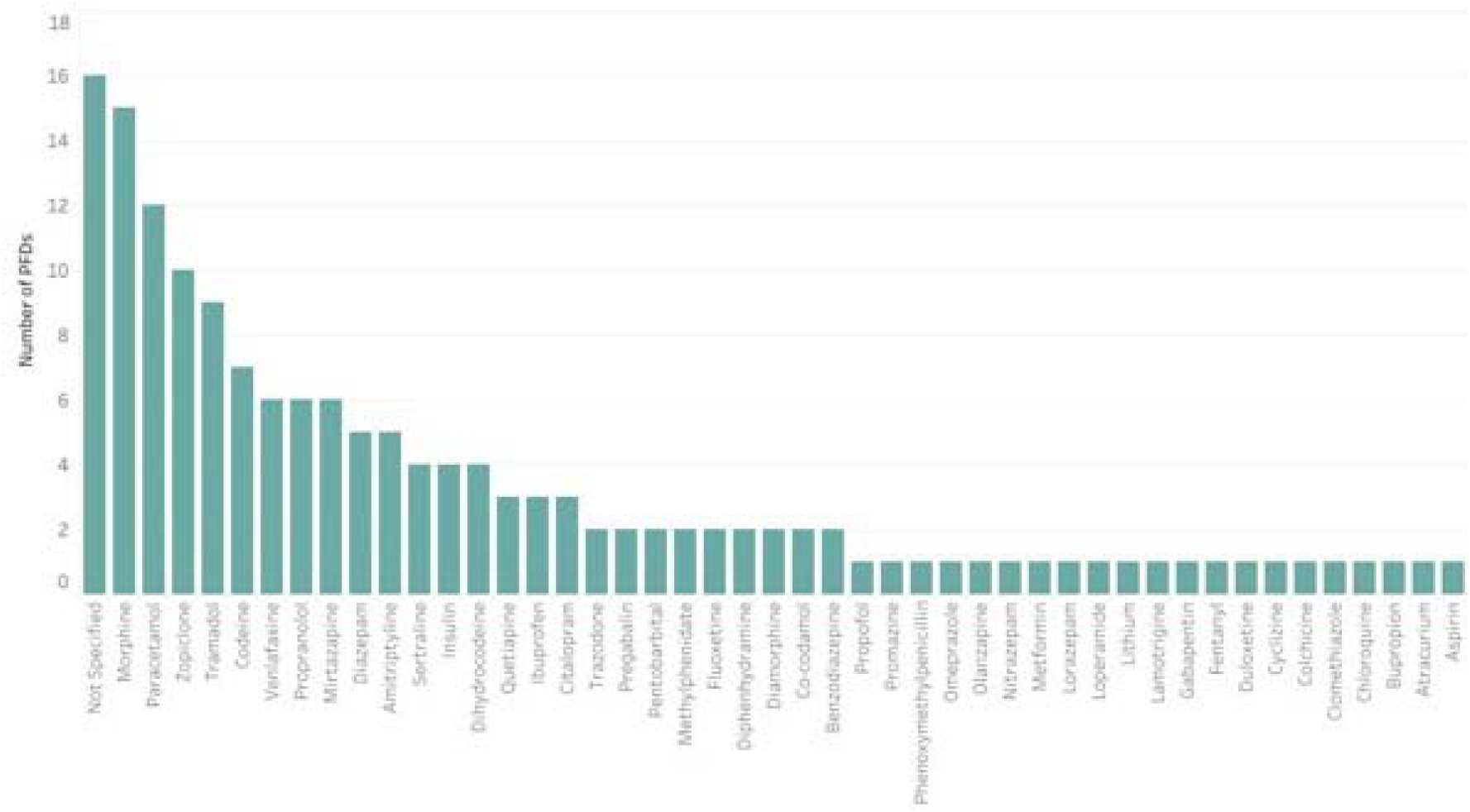
Types of medicines reported by coroners in suicide-related Prevention of Future Death reports (PFDs) in England and Wales between July 2013 and December 2019.

Across the 100 suicides involving medicines, there was an equal sex ratio of males (n=50) and females (n=50). The median age of the deceased was 42 years (IQR: 28-50 years; n=61). Depression (n=29) was the most common mental health problem reported by coroners, followed by anxiety disorders (n=12), emotionally unstable personality disorder/borderline personality disorder (n=11) and psychotic symptoms (n=10) (Supplement Figure S4).

Fifty-three coroners across 47 coroner areas wrote the 100 PFDs. Coroners in the Northwest (22%) and Southwest (16%) wrote the most, whereas coroners in the Northeast (3.0%) and Yorkshire and the Humber (3.0%) regions wrote the fewest (Figure 3). Manchester South issued the highest number of PFDs involving medicines and suicide (8.0%) (Supplement Table S3).

**Figure 3:**
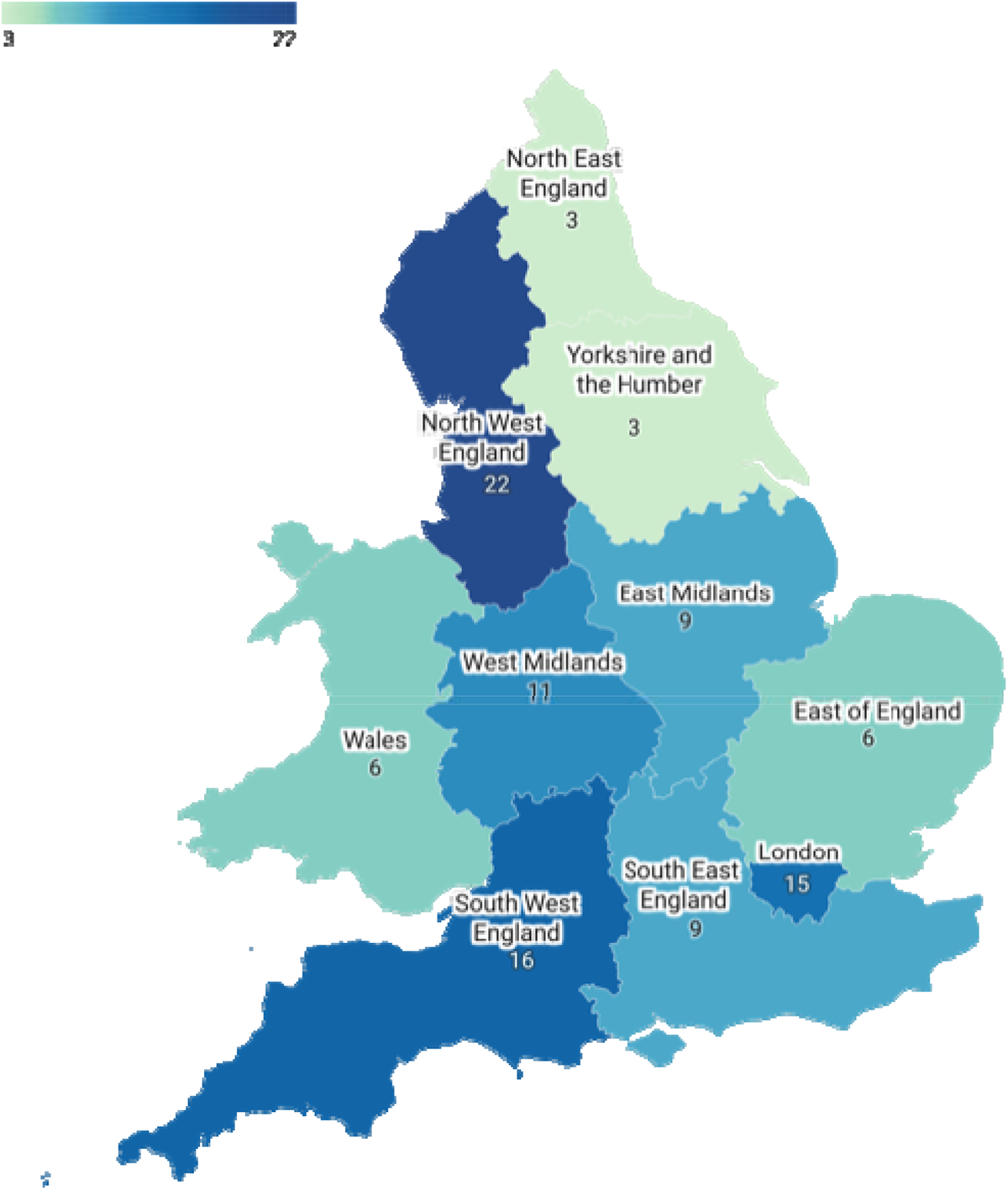
Choropleth map of medicine-related suicides reported in coroners’ Prevention of Future Deaths reports (PFDs) published in England and Wales from July 2013 to December 2019.

237 concerns were raised by coroners in the 100 PFDs. These were categorised using content analysis, which identified 18 themes (Figure 4). The concerns most frequently reported included procedural inadequacies (14%, n=32), inadequate documentation and communication (9.3%, n=22), inappropriate prescription access (8.9%, n=21), and improper risk assessment (7.2%, n=17; Supplement Table S4).

**Figure 4:**
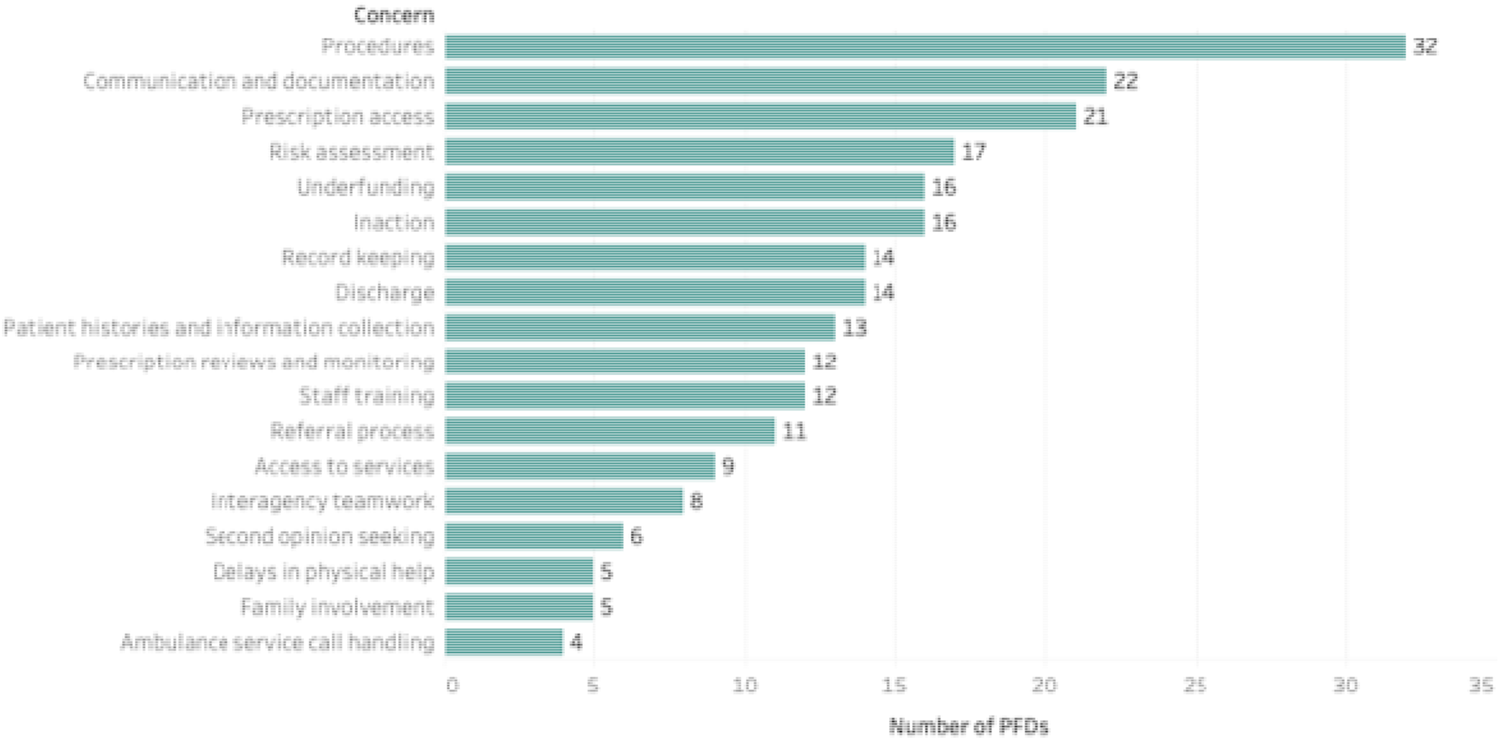
Concerns reported by coroners in 100 suicide-related Prevention of Future Deaths report (PFDs) involving medicines in England and Wales published between July 2013 and December 2019.

Coroners sent 203 PFDs to 145 individuals or organisations (Supplement Table S5). NHS trusts (31%; n=63), clinical commission groups (CCGs; 9.9%; n=20), and general practices (9.9%; n=20) were sent the most PFDs (Supplement Figure S5). By law, recipients must respond to the coroner within 56 days, but only 42% (n=85) were received within that time. Thirteen percent (n=27) were late, and 42% (n=86) were overdue. Government bodies (63%) and NHS bodies (62%) had the highest response rates, while law enforcement (44%) had the lowest response rates.

## Discussion

Coroners reported 100 deaths in PFDs that involved suicides and medicines before the Covid-19 pandemic. Coroners often raised concerns regarding procedural inadequacies, inadequate documentation and communication, and inappropriate prescription access. Opioids, antidepressants, and paracetamol were the most common medicines involved in suicide-related PFDs. NHS organisations and Government bodies received the most PFDs, but only 58% responded to the coroner.

### Comparison with previous research

A case series of all PFDs involving medicines similarly found that opioids and antidepressants were the most common medicines reported in PFDs (18). The ONS previously reported that opioids, antidepressants, and paracetamol were the most common drugs reported in self-poisoning suicides (31). The 2023 Healthcare Quality Improvement Partnership (HQIP) annual National Confidential Inquiry into Suicide and Safety in Mental Health also found that opioids were the most common type of drugs involved in suicides, most often obtained by prescription (32). An analysis of PFDs between January 2021 and October 2022 by the ONS found 164 PFDs categorised as suicide from the Judiciary website, which identified similar coroners’ concerns to our study (19). NHS Resolution has also reported similar concerns highlighted in suicide-related PFDs between 2015 and 2017, identifying issues in communication, risk assessments and the observational processes (13).

Despite the statutory requirement that mandates a response to the coroner’s PFD within 56 days (11), there is no mechanism to follow up responses or the ability to assess and ensure that actions proposed in responses are implemented. We found that over 40% of responses were still outstanding, making it challenging to understand whether action has been taken to reduce future deaths. Others have similarly found low response rates to PFDs (18, 33, 34). Future research should further examine responses and assess the factors that affect the response rate, such as the nature of death or the types of recipients.

We identified significant variation in the writing of PFDs across England and Wales. Others have similarly reported this (14, 18, 33). Future research should determine the factors driving this variation, including population statistics, mortality rates and coroners’ attitudes towards writing PFDs.

Male suicides account for three-quarters of suicides in England and Wales, leading to increased initiatives to reduce male-related suicide (2). However, we found no sex differences for medicines-related suicides. Studies have further highlighted the fact that female deaths are less likely to be reported to the coroner, be taken to inquest, and reach a conclusion of suicide, with accident/ misadventure and narrative verdicts being favoured conclusions for females (2, 27, 28, 35–37). Future research should account for potential sex biases in the writing of PFDs and the reporting of such deaths in PFDs.

### Strengths and limitations

This study is the first to have assessed medicines-related suicides reported in PFDs before the start of the Covid-19 pandemic. Our reproducible data collection methods allowed us to examine six and a half years of data from over 3000 PFDs. Previous studies have used less than two years of data and manually identify PFDs using the Judiciary website categorisation of deaths (14, 19, 33, 34). Of the 100 medicine-suicide PFDs in our study, only 19 were correctly categorised as ‘suicide’ on the Judiciary website, with most (n=48) categorised as ‘hospital-related’ (Figure S3 Supplement). Therefore, previous studies that used the Judiciary categorisation have underestimated the total number of deaths and have potential selection biases (14, 19, 33, 34). To be inclusive and reduce the coronial variation in concluding a suicide, we expanded previous definitions of suicides.

The 100 deaths included in this study do not represent all deaths deemed preventable involving suicides and medicines between July 2013 and December 2019, as the writing of PFDs depends on the coroner and the evidence presented during the death investigation. A qualitative interview of 32 senior coroners in 2009 showed that only 50% of respondents regarded public health as a key aspect of their duties (39). We acknowledge that other medicines, such as isotretinoin, are involved in preventable suicides, but we were limited by the information reported by coroners, with 16 PFDs not specifying the types of medicines involved (40). We encountered missing data for other essential variables, including age (not reported in 39% of PFDs), where the medicine was obtained (not reported in 47%), and the location of the suicide (not reported in 19%). Finally, it may also be possible that some PFDs and their responses may not have been published on the Judiciary website, resulting in publication and reporting biases.

### Implications

Suicide rates in England and Wales decreased from the early 1980’s to the early 2000s, in recent years rates have started to climb; in 2019 a suicide rate of 16.9 deaths per 100,000 in males was recorded, the highest since 2000; the female suicide rate in 2019 was equally the highest since 2004 (2, 41).

Concerns raised by coroners in PFDs should aid in suicide prevention and patient care. The ‘*10 ways to improve safety*’ by the *National Confidential Inquiry into Suicide and Safety in Mental Health* featured recommendations such as reducing drug and alcohol misuse, safer wards, and personalised risk management (32). Our study highlights the continued need for safer prescribing of opioids and antidepressants. In England, the prescribing of opioids and antidepressants has increased by 34% and 100% respectively between 2008 and 2018 (42, 43). The poor application of policies and procedures that were repeatedly identified by coroners should be addressed when updating NICE guidance on depression, which will be renewed in 2023 (44, 45). Medicines implicated in PFD suicides may be emerging evidence of risks associated with these medications. For example, the MHRA are considering if further action is required following concerns raised regarding increased risk of suicide when using ciprofloxacin and quinolones (46, 47).

In October 2023 the government will instate a new Health Services Safety Investigations Body, tasked with investigating mental health inpatient care settings, PFD reports could play a role in “how providers’…learn from tragic deaths that take place in their care” (48).

Our study examined suicide-related PFDs published before the Covid-19 pandemic (July 2013 to December 2019). There is substantial evidence that the Covid-19 pandemic negatively affected the mental health of many people, particularly the young, minorities, and women (49–53). The global burden of major depressive and anxiety disorders rose by 27% and 26% retrospectively during the Covid-19 pandemic (54). An analysis of PFDs during the Covid-19 pandemic (2021–2022) has since been conducted by the ONS, which found 164 suicide-related PFDs (19). In response to ONS’s analysis of suicide-related PFDs, the Samaritans stated that *“It’s hugely concerning that these common themes are consistently emerging and there’s no evidence to suggest this will improve without urgent, deliberate action.”*(55) Prescriptions for mental health conditions have been predicted to increase following the pandemic, and lockdowns may have increased the severity of mental illness (56, 57). Therefore, concerns raised by coroners in PFDs should be urgently used by policy makers to mitigate the increased risk of suicide.

Wider dissemination of PFDs and their responses could contribute to national initiatives to reduce suicides, as detailed by the NHS Mental Health Implementation Plan (58), national charities, and NHS Resolution (13, 55, 59). Despite the legal requirement under Regulation 29 of The Coroners (Investigations) Regulations 2013 to respond to coroners within 56 days (11), 42% of responses remained overdue. NHS Resolution recommended in 2018 that neighbouring NHS trusts should share learning from their PFDs and their responses to them, highlighting good practice to be regular meetings with their local and area coroner, to identify common themes arising (13). The Justice Select Committee’s report on the Coroner’s Service also acknowledged that the absence of follow-up on coroners’ PFDs is a missed opportunity and recommended that an Inspectorate be established to follow up on actions proposed in PFDs (60). The Government responded, stating that they were “not in a position to accept the recommendation at this stage” and that the cost would be disproportionate to the benefit it may bring (61). We therefore created a method, database, and resource (https://preventabledeathstracker.net/) to disseminate lessons from PFDs.

Future research should investigate the barriers and facilitators of writing and responding to PFDs, the suitability of concerns reported in suicide-medicine PFDs, the suitability of recipients of such PFDs, and the actions taken and proposed in the 117 responses received for suicide-medicine PFDs. Others can also use our data to analyse the 635 suicide-related PFDs that did not involve medicines between July 2013 and December 2019. The wide variation and missing data in PFDs also highlight areas for coronial training and the need to standardise reporting in PFDs and PFD-related research.

## Conclusions

PFDs hold a wealth of information regarding gaps in suicide prevention measures and the risks of medicine-related suicides. However, it is unclear if these PFDs are being used to benefit patient care and prevent future deaths. To aid in suicide prevention, PFDs should be disseminated more widely, responses to PFDs should be audited, and regular analyses of the spatial and temporal changes in concerns should be conducted. This study provides a tool (https://preventabledeathstracker.net/) to increase lessons learnt from coroners’ PFDs.

## Acknowledgements

not appliable

## Author Contributions

GA proposed the topic for research, wrote the study protocol and devised the algorithm for the eligibility of cases, screened the database for eligible case reports, extracted and analysed the data, and wrote the first draft of the manuscript. GCR designed the study, guided the writing of the study protocol, sourced the data, contributed to the web scraper, provided data management, dual screened the database for eligible cases, analysed the data, provided primary supervisory support and substantially contributed to subsequent manuscript drafts. JKA, RB, and CH reviewed the protocol and preliminary findings and provided supervisory support. All authors have full access to all the data in the study, read and approved the final manuscript, and accept responsibility to submit for publication.

## Transparency declaration

GA and GCR affirm that the manuscript is an honest, accurate, and transparent account of the study being reported; that no important aspects of the study have been omitted; and that any discrepancies from the study as planned have been explained.

The data that support the findings of this study are openly available in the OSF at https://doi.org/10.17605/OSF.IO/AD4UP

## Analytic code availability

The analyses in this study were descriptive in nature and therefore were conducted using the formulas in Google Sheets, which are all openly available in the OSF repository at https://doi.org/10.17605/OSF.IO/AD4UP

## Research Material availability

The research materials used in this study are openly available in the OSF at https://doi.org/10.17605/OSF.IO/AD4UP

## Ethical approval

The case reports have been publicly available on the Courts and Tribunals Judiciary website since 2013. Since the information is all publicly available, approval from an ethics committee was not required.

