## Supplementary Figure S1 for "Preventable suicides involving medicines before the covid-19 pandemic: a systematic case series of coroners’ reports in England and Wales"

**Figure S1:** Algorithm for selection of medicine-related Suicide PFDs

Yes

Does the coroner conclude:

Suicide, killed themselves, took own life

Was the death caused or contributed to by: self-hanging, strangulation and suffocation; self-harm; overdose; drowning; poisoning; fall and fracture; jumping or lying in front of a moving object; sharp object injury with

-

indications of intent

was the death caused or contributed to by: self-hanging, strangulation and suffocation; self-harm; overdose; drowning; poisoning; fall and fracture; jumping or lying in front of a moving object; sharp object injury

-

with undetermined intent or open conclusion

was the death caused or contributed to by: self-hanging, strangulation and suffocation; self-harm; overdose; drowning; poisoning; fall and fracture; jumping or lying in front of a moving object; sharp object injury

-

with no discussion of intent or the use of ‘undetermined intent’- where there is a discussion of mental health issues

Is this report a duplication?

was the death caused or contributed to by: self-hanging, strangulation and suffocation; self-harm; overdose; drowning; poisoning; fall and fracture; jumping or lying in front of a moving object; sharp object injury

-

attributed to as an accident where there is a history of mental health issues

Exclude

Yes

No

No

No

No

No

Yes

Yes

Yes

was this death caused or contributed to by acute onset of behavioural disturbance due to psychoactive drugs?

OR

Drug Overdose where individual has abstained resulting in reduced tolerance

did death occur on a railway line when individual accidentally trespassed on railway line while under the influence

Include

Exclude

Yes

Yes

Yes

No

No

No

did a medicine cause or contribute to the death?

No

Yes

**Figure S2.** Flow diagram of the screening of coroners’ Prevention of Future Deaths reports (PFDs) assessed in this study.

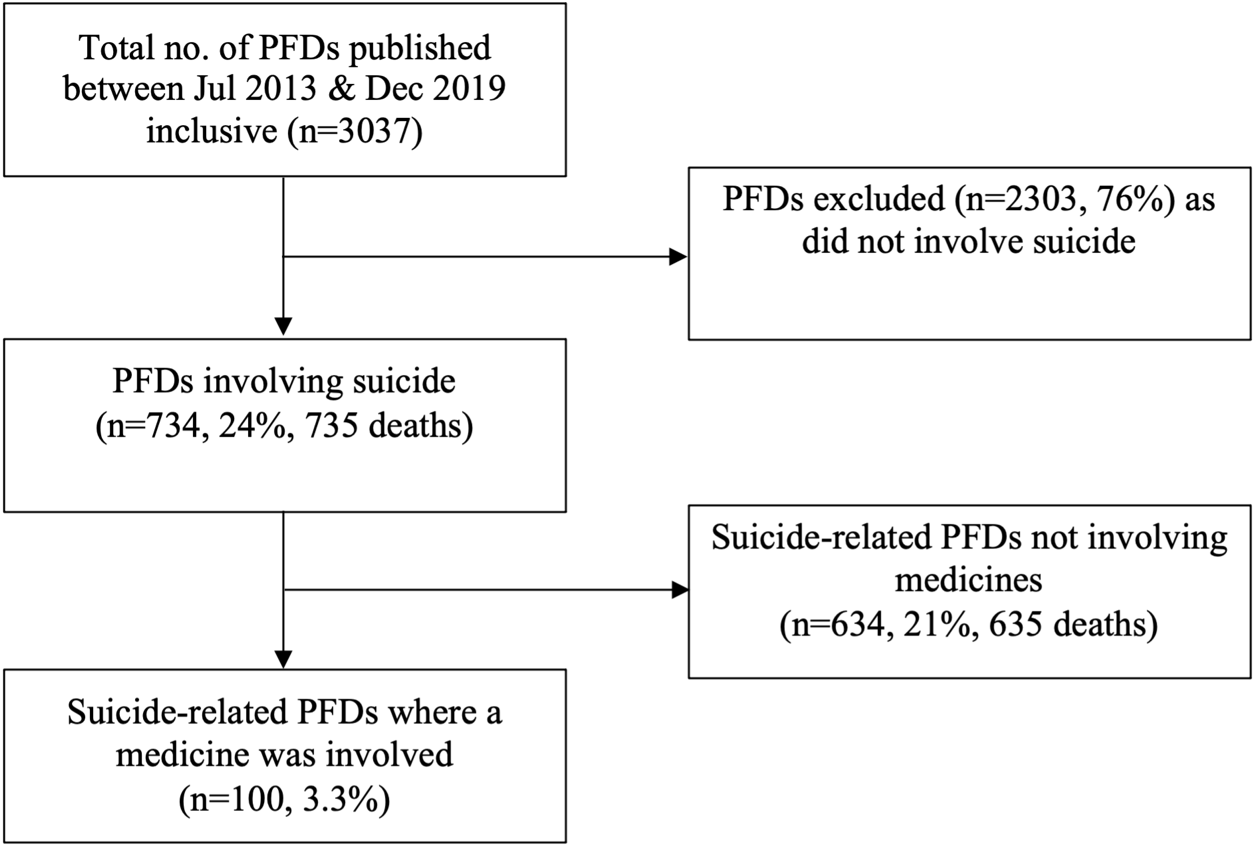

**Table S1:** Suicide type Prevent Future Death reports in comparison with the Office for National Statistics mortality data on suicides

| **Year** | **Total no. of PFDs** | **No. of Suicide PFDs** | **Medicine Suicides**  **(% of total)**  ***(%of Suicides*)** | **No. of Suicides reported to the ONS** | **% of Suicide PFDs vs ONS suicides** | ***% medicine suicide PFDs vs ONS Suicides*** |
| --- | --- | --- | --- | --- | --- | --- |
| 2013 | 173 | 38 | 4 (2.3)(*10.5*) | 5145 | 0.74% | 0.08% |
| 2014 | 555 | 120 | 19 (3.4)(*15.8*) | 5158 | 2.33% | 0.37% |
| 2015 | 478 | 99 | 10 (2.1)(*10.1*) | 5199 | 1.90% | 0.19% |
| 2016 | 469 | 118 | 10 (2.1)(*8.5*) | 4941 | 2.39% | 0.20% |
| 2017 | 436 | 104 | 21 (4.8)(*20.2*) | 4840 | 2.15% | 0.43% |
| 2018 | 412 | 118 | 17 (4.1)(*14.4*) | 5420 | 2.18% | 0.31% |
| 2019 | 514 | 137 | 19 (3.7)(*13.9*) | 5691 | 2.41% | 0.33% |
| **Total** | 3037 | 734 | 100 | 36394 | 2.02% | 0.27% |
| **Median (IQR)** | 469 (424-496) | 118 (101.5-119) | 17 (10-19) | 5158 (5043-5309.5) | 2.18 (2.03-2.36) | 0.31(0.2-0.35) |

PFDs: Preventions of future death reports; IQR: inter quartile range; ONS: Office for National Statistics

**Figure S3:** Judicary website categorisations in coroners’ Prevention of Future Deaths reports (PFDs) involving medicine-related suicides in England and Wales between July 2013 and December 2019**.** Categories featuring (2015) include reports from the year 2015 and onwards. PFDs can be categorised into one or more categories.
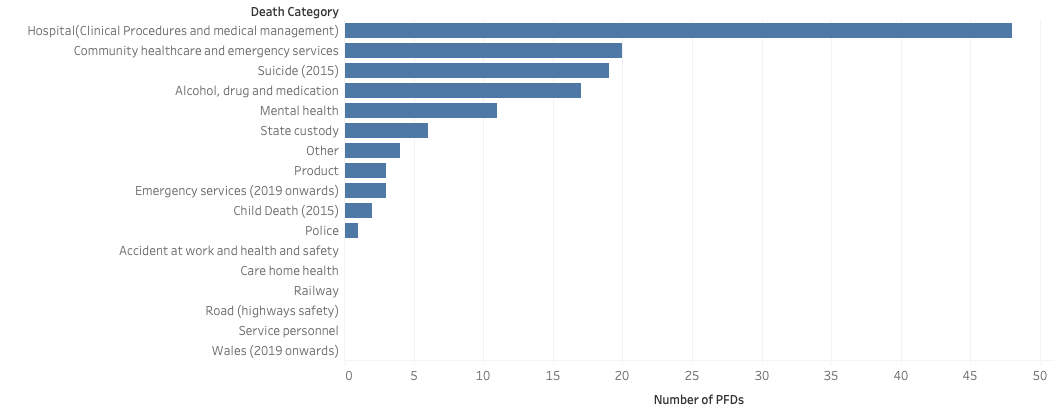

**Table S2**: Summary of the types of medicines reported in coroners’ Prevention of Future Deaths reports in England and Wales between July 2013 and December 2019, classified using the British National Formulary (BNF) and ordered descending.

| **Drug class** | **Drug Name** | **Number of PFDs** |
| --- | --- | --- |
| **Opioids** |  | ***40*** |
|  | Morphine | 15 |
|  | Tramadol | 9 |
|  | Codeine | 7 |
|  | Dihydrocodeine | 4 |
|  | Co-codamol | 2 |
|  | Diamorphine | 2 |
|  | Fentanyl | 1 |
| **Anti-depressants** |  | ***30*** |
| *Selective serotonin reuptake inhibitors* | | ***9*** |
|  | Sertraline | 4 |
|  | Citalopram | 3 |
|  | Fluoxetine | 2 |
| *Serotonin and norepinephrine reuptake inhibitors* | | ***8*** |
|  | Venlafaxine | 6 |
|  | Duloxetine | 1 |
|  | Bupropion | 1 |
| *Tri-cyclics* |  | ***7*** |
|  | Amitriptyline | 5 |
|  | Trazodone | 2 |
| *Tetra-cyclics* |  | ***6*** |
|  | Mirtazapine | 6 |
| **Non-opioid analgesics** |  | ***12*** |
|  | Paracetamol | 12 |
| **Non-benzodiazepine hypnotics and sedatives** | | ***11*** |
|  | Zopiclone | 10 |
|  | Clomethiazole | 1 |
| **Benzodiazepines** |  | ***7*** |
|  | Diazepam | 5 |
|  | Nitrazepam | 1 |
|  | Lorazepam | 1 |
| **Beta blocking agents, non-selective** |  | ***6*** |
|  | Propranolol | 6 |
| **Anti-epileptics** |  | ***4*** |
|  | Pregabalin | 2 |
|  | Lamotrigine | 1 |
|  | Gabapentin | 1 |
| **Insulin** |  | ***4*** |
| **Non-steroidal anti-inflammatories** | | ***4*** |
|  | Ibuprofen | 3 |
|  | Aspirin | 1 |
| **Second generation antipsychotics** | | ***4*** |
|  | Quetiapine | 3 |
|  | Olanzapine | 1 |
| **Barbiturates** |  | ***2*** |
|  | Pentobarbital | 2 |
| **Centrally acting sympathomimetics** | | ***2*** |
|  | Methylphenidate | 2 |
| **Intravenous anaesthetics** |  | ***1*** |
|  | Propofol | 1 |
| **First generation antipsychotics** | | ***1*** |
|  | Promazine | 1 |
| **Penicillins, beta-lactamase sensitive** | | ***1*** |
|  | Phenoxymethylpenicillin | 1 |
| **Biguanides** |  | ***1*** |
|  | Metformin | 1 |
| **Lithium salts** |  | ***1*** |
| **Plant alkaloids** |  | ***1*** |
|  | Colchicine | 1 |
| **Antimalarials** |  | ***1*** |
|  | Chloroquine | 1 |
| **Anti-propulsives** |  | ***1*** |
|  | Loperamide | 1 |
| **Neuromuscular blocking drugs, non-depolarising** | | ***1*** |
|  | Atracurium | 1 |
| **Proton pump inhibitors** |  | ***1*** |
|  | Omeprazole | 1 |

**Figure S4**: Mental health conditions reported in coroners Prevention of Future Deaths reports (PFDs) involving medicine-related suicides in England and Wales between July 2013 and December 2019. Where ‘not stated’: mental health disorder was mentioned but not specified; EUPD, BPD: Emotionally Unstable Personality Disorder, Borderline Personality Disorder; OCD: Obsessive Compulsive Disorder; ASD: Autism Spectrum Disorder; ADHD: Attention Deficit Hyperactivity Disorder; PTSD: Post-Traumatic Stress Disorder.

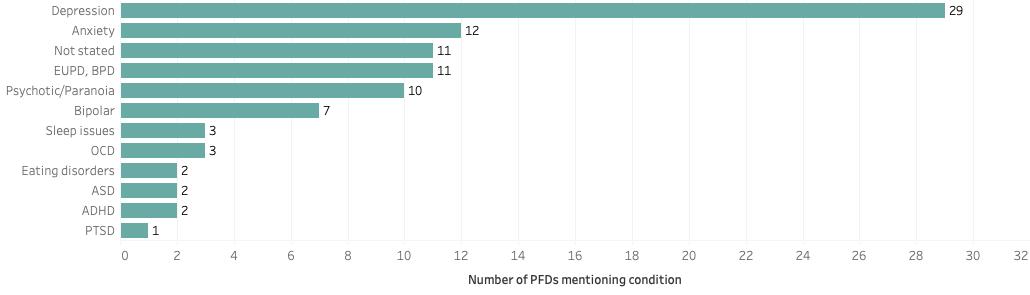

**Table S3**: Coroner areas where the 100 suicide-related Prevention of Future Deaths reports (PFDs) involving a medicine were written in England and Wales between July 2013 and December 2019 in descending order.

| **Region** | **No. of Cases %** |
| --- | --- |
| **North West England** | **22** |
| Cheshire | 1 |
| Cumbria (2015)* | 0 |
| South and East Cumbria^ | 0 |
| North and West Cumbria^ | 0 |
| Manchester city | 1 |
| Manchester North | 6 |
| Manchester South | 8 |
| Manchester West | 3 |
| Lancashire and Blackburn with Darwen (2017)* | 0 |
| Blackburn, Hyndburn and Ribble Valley^ | 0 |
| East Lancashire^ | 0 |
| Preston and West Lancashire^ | 0 |
| Blackpool/Fylde | 2 |
| Sefton, Knowsley and St Helens | 0 |
| Liverpool and Wirral (2015)* | 0 |
| Liverpool^ | 1 |
| Wirral^ | 0 |
| **South West England** | **16** |
| Avon | 0 |
| Cornwall and Isles of Scilly (2016)* | 3 |
| Cornwall^ | 0 |
| Isles of Scilly^ | 0 |
| Exeter and Greater Devon | 6 |
| Plymouth, Torbay and South Devon | 1 |
| Dorset | 1 |
| Gloucestershire | 1 |
| Somerset (2016)* | 0 |
| Eastern Somerset^ | 0 |
| Western Somerset^ | 0 |
| Wiltshire and Swindon | 4 |
| **London** | **15** |
| City of London | 1 |
| East London | 2 |
| Inner North London | 4 |
| Inner South London | 1 |
| Inner West London | 2 |
| North London | 1 |
| South London | 2 |
| West London | 2 |
| **West Midlands** | **11** |
| Herefordshire | 0 |
| Shropshire, Telford and Wrekin | 0 |
| Staffordshire South | 1 |
| Stoke-on-Trent and North Staffordshire | 0 |
| Warwickshire | 0 |
| Birmingham and Solihull | 6 |
| Black Country | 2 |
| Coventry | 2 |
| Worcestershire | 0 |
| **East Midlands** | **9** |
| Derby and Derbyshire | 1 |
| Leicester City and South Leicestershire | 2 |
| Rutland and North Leicestershire | 0 |
| Lincolnshire (2017)* | 1 |
| Central Lincolnshire^ | 1 |
| South Lincolnshire^ | 0 |
| Northamptonshire | 0 |
| Nottinghamshire | 4 |
| **South East England** | **9** |
| Berkshire | 0 |
| Brighton and Hove | 2 |
| Buckinghamshire | 1 |
| East Sussex | 0 |
| Central Hampshire | 1 |
| North East Hampshire | 0 |
| Portsmouth and South East Hampshire | 0 |
| Southampton and New Forest | 0 |
| Isle of Wight | 0 |
| Central and South East Kent | 0 |
| Mid Kent and Medway | 1 |
| North East Kent | 0 |
| North West Kent | 0 |
| Milton Keynes | 0 |
| Oxfordshire | 1 |
| Surrey | 1 |
| West Sussex | 2 |
| **East of England** | **6** |
| Bedfordshire and Luton | 0 |
| Cambridgeshire and Peterborough (2015)* | 0 |
| North and East Cambridgeshire^ | 0 |
| South and West Cambridgeshire^ | 0 |
| Peterborough^ | 0 |
| Essex | 0 |
| Hertfordshire | 0 |
| Norfolk | 4 |
| Suffolk | 2 |
| **Wales** | **6** |
| South Wales Central (2016)* | 3 |
| Powys, Bridgend and Glamorgan Valleys^ | 0 |
| Cardiff and Vale of Glamorgan^ | 0 |
| Carmarthenshire and Pembrokeshire | 0 |
| North Wales (East and Central) | 2 |
| Ceredigion | 0 |
| Gwent | 0 |
| Swansea and Neath Port Talbot | 1 |
| North West Wales | 0 |
| **Yorkshire and the Humber** | **3** |
| East Riding and Hull | 1 |
| North Lincolnshire and Grimsby | 0 |
| York City | 0 |
| North Yorkshire Eastern District | 0 |
| North Yorkshire Western District | 0 |
| South Yorkshire Eastern District | 0 |
| South Yorkshire Western District | 1 |
| West Yorkshire Eastern District | 0 |
| West Yorkshire Western District | 1 |
| **North East England** | **3** |
| County Durham and Darlington | 2 |
| Hartlepool and Teeside (2018)* | 0 |
| Teeside^ | 0 |
| Hartlepool^ | 0 |
| North Northumberland | 0 |
| South Northumberland | 0 |
| Gateshead and South Tyneside | 1 |
| Newcastle Upon Tyne | 0 |
| North Tyneside | 0 |
| Sunderland | 0 |

^ indicates a defunct coroner area that has been merged into a larger area; * indicates the new coroner areas with the year of the merger.

**Table S4.** Categorisation of concerns reported by coroners in the 100 suicide-related Prevention of Future Deaths reports (PFDs) involving a medicine written in England and Wales between July 2013 and December 2019, in descending order.

| **Theme** | **Concerns included** | **Number of PFDs including theme (% of 237 concerns)** |
| --- | --- | --- |
| Procedures | No protocol or procedures in place | 32 (13.5) |
|  | Inadequate procedures |  |
|  | Procedures were not understood by staff |  |
|  | Procedures were not followed |  |
|  | Observation procedures inadequate or not followed |  |
| Communication and documentation | Lack of transfer of information and documents | 22(9.3) |
|  | Information not transferred at discharge |  |
|  | Agencies not able to access each other’s records |  |
|  | Miscommunication between staff |  |
| Prescription Access | Prescriptions not secured in healthcare settings | 21(8.9) |
|  | Prescription too large |  |
|  | Prescription inappropriate in combination with pre-existing prescriptions |  |
|  | Prescriptions accessed at work with inadequate safety measures |  |
|  | Prescriptions obtained from family members |  |
|  | Prescriptions obtained from the internet or dark web |  |
|  | Inadequate regulation of medicine access |  |
| Risk assessment | No risk assessments carried out | 17(7.2) |
|  | Unclear risk assessment process |  |
|  | Failure to appreciate risks |  |
|  | No management of risks |  |
| Underfunding | Lack of hospital beds | 16(6.8) |
|  | Long waiting times for services |  |
|  | Staff shortages due to commissioning |  |
|  | Large caseloads |  |
|  | Long ambulance turnover times |  |
| Inaction | Inadequate action taken | 16(6.8) |
|  | No action taken by organisation to improve practices |  |
|  | Insufficient serious incident review |  |
| Discharge | Discharged too early | 14(5.9) |
|  | Inappropriate leave given |  |
|  | No follow-up or lost to follow-up |  |
|  | Insufficient advice given to patient at discharge |  |
|  | Onus on patient for contacting services |  |
| Record keeping | No records kept | 14(5.9) |
|  | Inaccessible records |  |
|  | Out-of-date records |  |
|  | No care plan or care plan out-of-date |  |
| Patient histories & collection of information | Health professionals failed to obtain sufficient information when assessing or engaging with patients | 13(5.5) |
| Prescription Reviews and monitoring | Patients taking controlled drugs and drugs of dependence not monitored or reviewed | 12(5.1) |
|  | Computer systems did not alert to dangerous prescribing |  |
|  | inadequate checks in systems for the prevention of unsafe prescriptions |  |
| Staff training | Lack of staff training | 12(5.1) |
|  | New training required |  |
| Referral process | Not referring | 11(4.6) |
|  | Not understanding the referral process |  |
|  | Issues in transfer process |  |
| Access to services | Lack of access to specialist services | 9(3.8) |
|  | Service not provided in area |  |
|  | Lack of continuity of care |  |
|  | Substance misuse services not able to provide mental health support or vice versa |  |
| Interagency teamwork | Lack of team meetings | 8(3.4) |
|  | Lack of joint working at *Assessment, Care in Custody/Detention and Teamwork* reviews |  |
|  | Emergency services not collaborating |  |
| Second opinion seeking | Patients not encouraged to seek second opinions | 6(2.5) |
|  | Staff not requesting another opinion |  |
| Family involvement | Family unsure of how to raise concerns | 5(2.1) |
|  | Family felt not listened to when raising concerns |  |
| Delays in physical help | Delay in requesting ambulance attendance | 5(2.1) |
|  | Delay in commencing cardio-pulmonary resuscitation |  |
|  | Delay in transferring test results in hospitals |  |
| Ambulance service call handling | Inappropriate categorisation to category 3 in emergency services call handling | 4(1.7) |
|  | Lack of follow-up welfare calls |  |

**Table S5.** Recipients sent coroners’ Prevention of Future Death reports (PFDs) involving medicine-related suicides in England and Wales published between July 2013 and December 2019. Responses were classified as early (delivered before 56 days), on-time (delivered on the 56th day), late (submitted after 56 days), or overdue (when no response was present on the Judiciary website).

| **Addressee** | **No. of reports received** | **No. of responses** | **Response rate (%)** |  |  |  |  |  |
| --- | --- | --- | --- | --- | --- | --- | --- | --- |
|  |  |  |  | **% Early** | **%On time** | **%Late** | **%Overdue** | **%Unclear** |
| **NHS Body** | **138** | **86** | **62** | **36** | **10** | **14** | **38** | **1** |
| CCGs | 20 | 13 | 65 | 45 | 5 | 10 | 35 | 5 |
| NHS addiction services | 1 | 0 | 0 | 0 | 0 | 0 | 100 | 0 |
| NHS England | 9 | 6 | 67 | 22 | 0 | 44 | 33 | 0 |
| NHS England south-west | 1 | 0 | 0 | 0 | 0 | 0 | 100 | 0 |
| NHS Trusts and University Health Boards | 63 | 40 | 63 | 33 | 14 | 14 | 38 | 2 |
| Ambulances | 8 | 4 | 50 | 38 | 0 | 13 | 50 | 0 |
| Hospitals | 9 | 5 | 56 | 22 | 33 | 0 | 44 | 0 |
| GP practices | 20 | 11 | 55 | 35 | 0 | 20 | 45 | 0 |
| Mental Health Trusts | 6 | 6 | 100 | 100 | 0 | 0 | 0 | 0 |
| NHS Pathways | 1 | 1 | 100 | 0 | 100 | 0 | 0 | 0 |
| **Government** | **24** | **15** | **63** | **33** | **0** | **25** | **38** | **4** |
| Local Authorities | 6 | 4 | 67 | 50 | 0 | 17 | 33 | 0 |
| DHSC | 11 | 8 | 73 | 27 | 0 | 45 | 27 | 0 |
| Home Office | 2 | 0 | 0 | 0 | 0 | 0 | 100 | 0 |
| Home Secretary | 1 | 0 | 0 | 0 | 0 | 0 | 100 | 0 |
| Secretary of State for Health | 1 | 1 | 100 | 100 | 0 | 0 | 0 | 0 |
| Welsh Minister for Health | 2 | 2 | 100 | 50 | 0 | 0 | 0 | 50 |
| Advisory Council on the misuse of drugs | 1 | 0 | 0 | 0 | 0 | 0 | 100 | 0 |
| **Law enforcement** | **18** | **8** | **44** | **22** | **11** | **11** | **56** | **0** |
| Police | 5 | 3 | 60 | 60 | 0 | 0 | 40 | 0 |
| MOJ | 2 | 1 | 50 | 0 | 0 | 50 | 50 | 0 |
| MOD | 1 | 1 | 100 | 100 | 0 | 0 | 0 | 0 |
| Mitie Care and Custody | 1 | 0 | 0 | 0 | 0 | 0 | 100 | 0 |
| Prisons | 3 | 0 | 0 | 0 | 0 | 0 | 100 | 0 |
| Director of public prosecutions | 1 | 0 | 0 | 0 | 0 | 0 | 100 | 0 |
| National offender management service | 1 | 0 | 0 | 0 | 0 | 0 | 100 | 0 |
| HM prison and probation service | 2 | 2 | 100 | 0 | 50 | 50 | 0 | 0 |
| National police chiefs’ council | 1 | 0 | 0 | 0 | 0 | 0 | 100 | 0 |
| G4S | 1 | 1 | 100 | 0 | 100 | 0 | 0 | 0 |
| **Professional bodies** | **13** | **7** | **54** | **31** | **8** | **0** | **46** | **15** |
| CQC | 1 | 1 | 100 | 0 | 0 | 0 | 0 | 100 |
| PHE | 1 | 1 | 100 | 100 | 0 | 0 | 0 | 0 |
| MHRA | 3 | 0 | 0 | 0 | 0 | 0 | 100 | 0 |
| GMC | 2 | 1 | 50 | 50 | 0 | 0 | 50 | 0 |
| GPC | 1 | 0 | 0 | 0 | 0 | 0 | 100 | 0 |
| RCGP | 2 | 2 | 100 | 100 | 0 | 0 | 0 | 0 |
| Toxbase | 1 | 1 | 100 | 0 | 0 | 0 | 0 | 100 |
| University bodies | 2 | 1 | 50 | 0 | 50 | 0 | 50 | 0 |
| **Other** | **10** | **2** | **20** | **20** | **0** | **0** | **80** | **0** |
| Housing support/associations | 4 | 2 | 50 | 50 | 0 | 0 | 50 | 0 |
| Private healthcare companies | 3 | 0 | 0 | 0 | 0 | 0 | 100 | 0 |
| Blank | 1 | 0 | 0 | 0 | 0 | 0 | 100 | 0 |
| Livewell Southwest | 1 | 0 | 0 | 0 | 0 | 0 | 100 | 0 |
| Post-prison support charity | 1 | 0 | 0 | 0 | 0 | 0 | 100 | 0 |

*CCG: Crown Commissioning Group; CQC: Care Quality Commission; DHSC: Department Of Health and Social Care; G4S: Group 4 Securicor; GMC: General Medical Council: GPC: General Pharmaceutical Council; MHRA: Medicines and Healthcare Products Regulatory Agency; MOD: Ministry Of Defence; MOJ: Ministry of Justice; PHE: Public Health England; RCGP: Royal College of General Practitioners.*

**Figure S5**

Recipients sent coroners’ Prevention of Future Death reports (PFDs), involving medicine-related suicides in England and Wales published between July 2013 and December 2019, categorised.

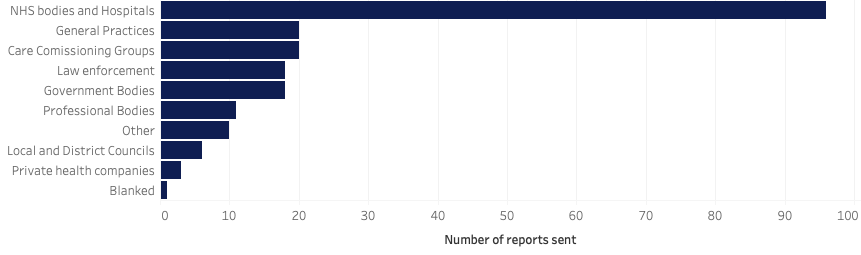
